# Musculoskeletal Health in Ageing Transgender and Gender Diverse Women: A Pilot Study

**DOI:** 10.64898/2025.12.08.25341839

**Authors:** Blair R. Hamilton, Nina Mitic, Charlotte E. Linscott, Ke Hu, Maaike Esselaar, Alex Ireland

**Affiliations:** Department of Sport and Exercise Sciences, Manchester Metropolitan University, Manchester, UK; Institute of Sport, Manchester Metropolitan University, Manchester, UK; The Tavistock and Portman NHS Foundation Trust, London, UK; Department of Sports and Health Sciences, Hong Kong Baptist University, Hong Kong SAR, China; Department of Life Sciences, Manchester Metropolitan University, Manchester, UK

## Abstract

**Introduction:** Age-related declines in bone and muscle health contribute substantially to frailty, falls, and functional loss in older adults. Despite the established musculoskeletal effects of hormone therapy (HT) in younger transgender and gender diverse women (TGDW), data on ageing TGDW do not exist. TGDW ≥50 years receiving feminising HT exhibit an 80% higher fracture incidence than cisgender adults. This pilot study explored the practicality of community-based recruitment of TGDW receiving HT and provided the first characterisation of musculoskeletal health in this population.

**Methods:** Cross-sectional pilot study recruited forty-eight TGDW aged ≥50 years undergoing feminising HT through two UK community festivals. Body composition (bioelectrical impedance analysis), bone quality (quantitative ultrasound of the radius and tibia), muscle strength and power (handgrip dynamometry, countermovement jump, and 5× sit-to-stand test), and physical activity (IPAQ-SF)were analysed using descriptive statistics, one-sample t-tests against clinical thresholds, and regression models exploring associations between HT duration and musculoskeletal outcomes, adjusting for age, BMI, and physical activity level.

**Results:** Sarcopenia incidence varied from 9% (handgrip strength) to 38% (countermovement jump power), and 38% of participants recorded counter-movement jump power below sarcopenia thresholds, whilst 21% could not complete the sit-to-stand test. Osteoporosis prevalence (8-13%) was higher than in the UK population (5%). Seventy-three per cent of participants were classified as overweight, and 33% classified as obese, higher than UK population norms (26%). Twenty-three per cent of TGDW were classed as physically active compared to their age-matched population (63%). Regression analyses showed no significant association between hormone therapy duration and musculoskeletal outcomes, whereas BMI (countermovement jump power β = 192.1, *p_adj_*=0.01; handgrip β = 0.60, *p_adj_*=0.02) was a stronger predictor of function.

**Conclusion:** High sarcopenia and osteoporosis prevalence, elevated obesity, and low physical activity highlight areas of concern. Larger longitudinal studies to characterise musculoskeletal ageing using clinical measures are needed.

**WHAT IS ALREADY KNOWN ON THIS TOPIC:**

- Ageing is associated with declines in bone and muscle health, leading to frailty, falls, and functional loss.
- Transgender and gender-diverse adults (TGDW) receiving feminising hormone therapy (HT) may experience musculoskeletal changes, but data on those aged ≥50 years are absent.

**WHAT THIS STUDY ADDS:**

- Provides the first characterisation of musculoskeletal health in ageing TGDW, highlighting osteoporosis and sarcopenia prevalence, substantial variation in bone quality and muscle function, functional limitations in a subset, elevated BMI and reduced physical activity.
- Demonstrates the feasibility of multi-domain musculoskeletal assessment in this population and identifies BMI and physical activity as stronger determinants of function than hormone therapy duration.

**HOW THIS STUDY MIGHT AFFECT RESEARCH, PRACTICE, OR POLICY:**

- Supports the need for larger longitudinal studies to track musculoskeletal ageing in TGDW.
- Highlights key modifiable risk factors (BMI, activity) for interventions, informing clinical management and health policy for older TGDW populations.

## Introduction

Musculoskeletal conditions represent the third-largest area of healthcare expenditure in the National Health Service, costing approximately £5 billion annually [1]. In the UK, the number of transgender and gender diverse adults increased fivefold from 2000-2018 with 262,000 individuals identifying as transgender and gender diverse in the 2021 Census [2]. Ninety-four per cent of all service users attending gender identity clinics in the UK receive gender-affirming hormone therapy (HT) to align their bodies with their gender identity [3]. Feminising HT exerts distinct effects on musculoskeletal health in transgender and gender diverse women (TGDW) [4, 5]. Among younger TGDW (mean age ≈ 30 years), HT has been associated with increased hip bone mineral density, fat mass, body mass index, and body fat percentage, alongside reductions in lean mass and thigh muscle area [6]. However, musculoskeletal health in *ageing* TGDW has never been characterised, despite evidence that TGDW aged over 50 years receiving feminising HT experience an 80% higher fracture incidence compared with cisgender adults [7].

Multiple lifestyle and behavioural factors may further compromise musculoskeletal integrity in this population. Lower levels of physical activity, greater social isolation, and reduced quality of life have been reported among TGDW [8], with only 24% exercising at least three times per week compared to 36–38% of cisgender adults [9]. Higher rates of smoking [10], alcohol consumption, and obesity [11] may also heighten vulnerability to musculoskeletal disease [12]. These risks mirror those observed in ageing adults generally, of whom approximately 60% are affected by sarcopenia [13] or osteoporosis [14], both linked to falls, frailty, and mortality.

Given the growing TGDW population [15], the dynamic physiological effects of HT, and the accumulation of age-related risk factors, characterising musculoskeletal health in older TGDW, is both timely and essential. This pilot study aimed to evaluate study feasibility and provide the first comprehensive characterisation of musculoskeletal health in ageing TGDW.

## Methods

### Participants

Forty-eight TGDW aged ≥50 years undergoing feminising hormone therapy (HT) were recruited through community festivals (Sparkle, Manchester, UK; TransPride, Brighton, UK). Inclusion criteria were: (1) aged 50 years or older; (2) on oestrogen-based HT (with or without anti-androgens) for at least 3 months; (3) able to provide written informed consent. Exclusion criteria included diagnosed metabolic bone disease (other than osteopenia/osteoporosis), recent fracture (<6 months), or medications known to substantially affect musculoskeletal outcomes (e.g., corticosteroids, bisphosphonates). Ethical approval was obtained from Manchester Metropolitan University [Ethos ID: 77000], and all participants provided written informed consent. This analysis was originally designed to sample both TGDW and transgender and gender diverse men. However, we were unable to recruit a sufficient number of participants undergoing masculinising HT to allow meaningful analysis (*n* = 3).

### Reference Data

All comparisons were made against reference values for individuals assigned male at birth, based on the hypothesis that this represents the musculoskeletal trajectory participants would have followed without HT. Age-matching was applied where possible. Reference data were sourced from established guidelines for sarcopenia [16], adiposity [17], and osteoporosis [18]. Clinical cut-offs for mobility limitation [19], sarcopenia [20], and adiposity [21] were obtained from peer-reviewed literature. Prevalence estimates were obtained from the Active Lives Survey (UK) for physical activity [22], National Health Service reports for obesity [17], the Scorecard for Osteoporosis in Europe and U.S. data for osteoporosis [23, 24], and the UK Biobank for sarcopenia [25].

### Body Composition

Body composition was assessed via bioelectrical impedance analysis (BIA) using the BodyStat 1500 device (Douglas, Isle of Man). Participants were tested in a supine position following a 5-minute rest, with electrodes placed at the right hand and foot according to manufacturer guidelines. The precision of BIA is 4.2%[26]

### Bone Health

Bone quality was evaluated using quantitative ultrasound (QUS) at the 50% of the radius (from Cubitus to tip of the Digitus Medius) and 50% tibia (Calcaneus to Tibial Tuberosity) with a Sunlight MiniOmni™ Bone Densitometer (BeamMed™, Florida, USA). Measures included speed of sound (SOS, m/s), T-scores and Z-scores. QUS precision was 2% at the radius and 5% at the tibia (Supplementary File CV QUS [27]

### Muscle Strength and Power

Handgrip strength was measured using a TAKEI 5401 dynamometer (Takei Scientific Instruments Co., Japan). The participant’s dominant hand was tested three times with 30s rest in between. The participant was seated on a chair with their right hand in lateral rotation, with the radius and ulna at a 90° angle to the humerus. The full methodology is described in detail in Hamilton (2024 [28]). The mean and maximum of three attempts were recorded.

Lower body power was assessed via countermovement jump (CMJ) on Hawkins Dynamic Force Plates (Westbrook, ME, USA). Participants performed three valid jumps (hands on hips, countermovement <45°), with mean jump height, peak propulsive force and power recorded. Countermovement jump was only assessed in Sparkle, Manchester, due to inclement weather. The portable Hawkins Dynamic Force Plates are a valid alternative to the industry gold standard in ground force plates, as fixed or proportional bias has not been identified for the variable of jump height [29]

Sit-to-Stand (STS) performance was measured in the 5-times STS format, recording total completion time (s). A standardised chair (45 cm) was used, and safety monitoring was performed throughout testing. Relative STSx5 mean power was calculated using a validated equation [30, 31]:

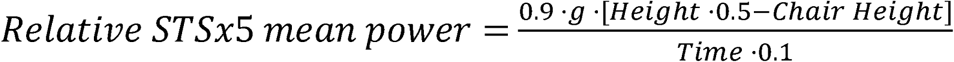

In this equation, *g* represents gravitational acceleration (9.81 m·s^⁻²^), and both body height and chair height are measured in metres (m). Time is recorded in seconds (s). The constants 0.9 and 0.1 are biomechanics-derived coefficients that account for movement efficiency and body segment proportions.

### Questionnaire

Physical activity levels were assessed using the International Physical Activity Questionnaire—Short Form (IPAQ-SF), which captures frequency and duration of walking, moderate, and vigorous activity, and sitting time over the previous 7 days. Data were cleaned and processed according to the IPAQ scoring protocol [32]. The time spent in each activity domain was multiplied by its corresponding MET value (walking = 3.3 METs; moderate = 4.0 METs; vigorous = 8.0 METs) and expressed as MET-minutes per week. These physical activity data were collected to adjust for potential lifestyle confounders.

### Statistical Analysis

All analyses were conducted in Jamovi (v2.7.6), with alpha values set at <0.05. Participant demographics, HT duration, and outcome variables were summarised using mean ± SD. An ANOVA compared musculoskeletal outcomes across these groups of HT duration: acute (<2 years), moderate (2–5 years), and long-term (>6 years). To quantify deviations from established musculoskeletal function cut-offs, we employed one-sample t-tests comparing sample means against recognised clinical thresholds. The null hypothesis was that participants met the definition of “normal” or “healthy” function, as specified by clinical guidelines (e.g., bone mineral density T-score > −2.5 indicating absence of osteoporosis; sit-to-stand (x5) < 15 s indicating preserved functional mobility). The alternative hypothesis was directional and reflected impairment, such that values beyond the cut-off indicated the presence of abnormal musculoskeletal status (e.g., HD: μ ≤ −2.5 for osteoporosis, or HD: μ ≥ 15 for sit-to-stand time). Effect sizes were estimated using Cohen’s d (standardised mean differences). Exploratory analyses examined associations between HT duration and musculoskeletal outcomes using linear regression, while multiple regression models adjusted for potential covariates, including age, BMI, and Metabolic Equivalent of Task (METS) minutes. Where covariates did not contribute significantly, reduced models are reported for parsimony. Model assumptions (normality, homoscedasticity, linearity, and absence of multicollinearity) were verified before interpretation. To control for false discovery rate, the Benjamini–Hochberg correction was applied within each outcome domain.

### Post-Hoc Power Analysis

Post hoc power analyses were performed using G*Power version 3.1.9.7 to determine the achieved statistical power for each hypothesis test. Analyses were conducted under the assumption of an α-level of 0.05, using the observed effect size (Cohen’s d) calculated from the sample mean difference and standard deviation, along with the actual sample size and degrees of freedom for each test. For each analysis, the achieved power (1 − β) is presented in Table 2. For tests that reached statistical significance, the critical t-value and non-centrality parameter are reported in Table 3 to indicate the sensitivity of the study to detect the observed effects.

### Prevalence

Prevalence was calculated as the proportion of participants meeting the specified clinical threshold, divided by the total number of participants assessed for that outcome, and expressed as a percentage.

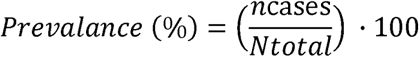

Where *n*_cases_ = number of participants meeting threshold and *N*_total_ = total number of participants assessed.

### Ethics Statement

This study was approved by the Manchester Metropolitan University Research Ethics Committee (Ethos ID: 77000) and conducted in accordance with the principles outlined in the Declaration of Helsinki [33]. All participants provided informed consent prior to participation.

## Results

### Participant Characteristics

The main characteristics of participants (*n* = 48) are shown in Table 1. Eighty-one per cent of participants consumed alcohol, 2% of participants took calcium, 35% took Vitamin C, and 42% took Vitamin D supplements.

Sarcopenia prevalence measured by hand grip and sit-to-stand performance was higher (9% and 21% respectively, Table 2) compared to that observed in UK normative data. Osteoporosis prevalence measured by QUS was higher (8% at the radius and 13% at the tibia, Table 2), while Obesity prevalence was also higher at 33% (Table 2) than UK normative data. TGDW were also less physically active (23% Table 2) than the UK population

The ANOVA comparing acute, moderate, and long-term HT duration groups revealed no significant differences for any musculoskeletal or functional measure (all p > 0.05), indicating that duration category did not meaningfully impact outcomes in this cohort.

**Table 1.**
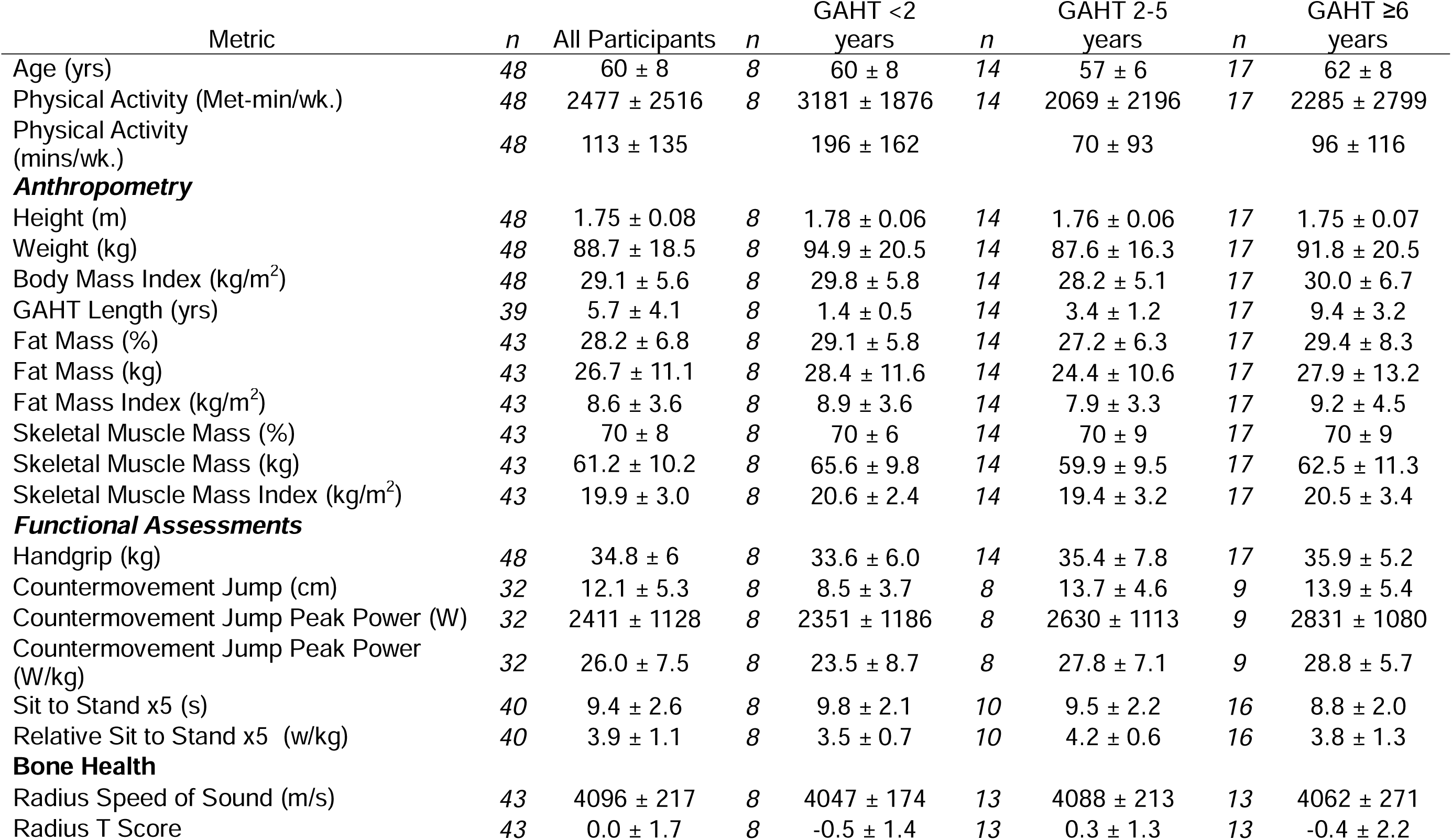
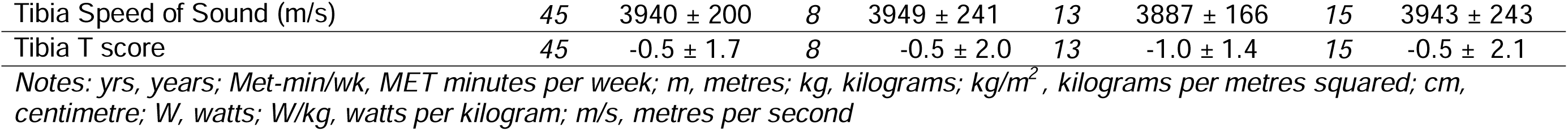
Participant Characteristics.

**Table 2.** One-Sample T-Test Analysis against Clinical and Proposed Musculoskeletal Health Outcomes.

| Metric | Cut-off | Statistic | df | p | Mean Difference | 95% Confidence Interval |  | Cohen's d<br>Effect Size | 95% Confidence Interval |  | Achieved Power |
| --- | --- | --- | --- | --- | --- | --- | --- | --- | --- | --- | --- |
|  |  |  |  |  |  | Lower | Upper |  | Lower | Upper |  |
| European Working Group on Sarcopenia in Older People 2 guidelines [16] |  |  |  |  |  |  |  |  |  |  |  |
| Hand Grip (kg) | < 27 | 8.9 | 47 | 1.00 | 7.8 | -inf | 9.7 | 1.3 | 0.9 | 1.7 | 1.00 |
| Prevalence | Sarcopenia | 9% |  |  | UK | 5% [25] |  |  |  |  |  |
| Sit to Stand (5s) | >15 | -13.5 | 39 | 1.00 | -5.6 | -6.31 | inf | -2.1 | -2.7 | -1.6 | 1.00 |
| Prevalence | Sarcopenia | 21% |  |  | UK | 5% [25] |  |  |  |  |  |
| Sarcopenia Cut-offs proposed by Hong et. al [20] |  |  |  |  |  |  |  |  |  |  |  |
| Countermovement Jump (W.kg) | <23.8 | 1.7 | 31 | 0.95 | 2.2 | -inf | 4.5 | 0.3 | -0.1 | 0.6 | 0.51 |
| Prevalence | Sarcopenia | 38% |  |  |  |  |  |  |  |  |  |
| Mobility limitations proposed by Alcazar et al [19] |  |  |  |  |  |  |  |  |  |  |  |
| Sit to Stand (5s [W.kg]) | <2.6 | 7.8 | 40 | 1.00 | 1.3 | -inf | 1.58 | 1.2 | 0.8 | 1.6 | 1.00 |
| Prevalence | Mobility Limited | 25% |  |  |  |  |  |  |  |  |  |
| World Health Organisation Standards for Bone Densitometry [18]. |  |  |  |  |  |  |  |  |  |  |  |
| Radius |  |  |  |  |  |  |  |  |  |  |  |
| Normal | > -1.0 | 4.0 | 42 | <.001 | 1.0 | 0.6 | inf | 0.6 | 0.3 | 0.9 | 0.99 |
| Osteoporosis | <-2.5 | 10.0 | 42 | 1.00 | 2.5 | -inf | 2.95 | 1.5 | 1.1 | 2.0 | 1.00 |
| Tibia |  |  |  |  |  |  |  |  |  |  |  |
| Normal | > -1.0 | 1.9 | 44 | 0.03 | 0.5 | 0.1 | inf | 0.3 | 0.0 | 0.6 | 0.63 |
| Osteoporosis | <-2.5 | 7.7 | 44 | 1.00 | 2.0 | -inf | 2.41 | 1.2 | 0.8 | 1.5 | 1.00 |
| Prevalence of Osteoporosis Vs Population Data [23, 24]. |  |  |  |  |  |  |  |  |  |  |  |
| Osteoporosis | Radius | 8% | Tibia | 13% | UK | 5% | Europe | 6% | USA | 6% |  |
| National Health Service Standards for Obesity [17] |  |  |  |  |  |  |  |  |  |  |  |
| Body Mass Index |  |  |  |  |  |  |  |  |  |  |  |
| Overweight | >25 | 5.1 | 47 | **<.001** | 4.1 | 2.74 | inf | 0.7 | 0.4 | 1.1 | 1.00 |
| Obese | >30 | -1.1 | 47 | 0.87 | -0.91 | -2.26 | inf | -0.2 | -0.5 | 0.1 | 0.39 |
| Prevalence | Overweight | 73% | UK | 64% | Obese | 33% | UK | 26% |  |  |  |
| Body Fat Percentage as proposed by Potter et al. [21] |  |  |  |  |  |  |  |  |  |  |  |
| Overweight | >25 | 3.1 | 42 | **0.002** | 3.2 | 1.49 | inf | 0.5 | 0.2 | 0.8 | 0.94 |
| Obese | >30 | -1.7 | 42 | 0.95 | -1.8 | -3.5 | inf | -0.3 | -0.6 | 0.4 | 0.61 |
| Prevalence | Overweight | 58% |  |  | Obese | 35% |  |  |  |  |  |
| Physical Activity Classification [22] |  |  |  |  |  |  |  |  |  |  |  |
| Physically Active (mins/wk) | >150 | -1.9 | 46 | 0.97 | -36.7 | -68.8 | inf | -0.3 | -0.6 | 0.2 | 0.64 |
| Fairly Active (mins/wk) | <149 | -1.8 | 46 | **0.04** | -35.7 | -inf | -2.5 | -0.3 | -0.6 | 0.3 | 0.65 |
| Physically inactive(mins/wk) | <30 | 4.2 | 46 | 1.00 | 83.4 | -inf | 116.5 | 0.6 | 0.3 | 0.9 | 0.99 |
| Prevalence | Inactive | 25% | Fairly Active | 52% | Physically Active | 23% | Physically Active | UK 55-64 yrs | 63% |  |  |

**Table 3.** Post-Hoc Statistical Power Analysis.

| **Outcome** | **Cut off** | **df** | **p** | **Critical t** | **Non-centrality** | **β**<br>**(Type II Error)** | **Power**<br>**Ranking** |
| --- | --- | --- | --- | --- | --- | --- | --- |
| Radius T score | Normal > -1.0 | 42 | **<.001** | 1.7 | 3.9 | 0.01 | **Excellent** |
| Tibia T score | Normal >-1.0 | 44 | 0.03 | 1.7 | 2.0 | 0.37 | Low |
| Body Mass Index (kg/m<sup>2</sup>) | Overweight >25 | 47 | **<.001** | 1.7 | 4.9 | 0.00 | **Excellent** |
| Body Fat (%) | Overweight >25 | 42 | **0.002** | 1.7 | 3.2 | 0.06 | **Excellent** |
| Physical Activity (mins/wk) | Fairly Active <149 | 46 | 0.04 | -1.7 | -2.1 | 0.35 | Low |

### Exploratory Linear Analyses

#### Anthropometry

Multiple linear regressions were conducted to examine whether age, MET-minutes, and hormone duration predicted fat mass, lean mass, BMI, and bone health (radius and tibia T-scores). None of the models were statistically significant (all p > 0.05), and the variance explained was low across outcomes (R² range: 0.03–0.16). No individual predictors reached significance.

### Functional Measurements

Handgrip and sit-to-stand models were not statistically significant (all p > 0.05), with no meaningful predictors identified.

### Countermovement Jump (CMJ)

The model was highly significant, F_(4, 19)_ = 41.64, p < 0.001, explaining 90% of the variance (R² = 0.90, adjusted R² = 0.88). BMI was a strong positive predictor (ß = 192.1, 95% CI 160.0–224.2, p < 0.001), and MET-minutes was a significant negative predictor (ß = −81.5 per 1,000 MET-minutes, 95% CI −150.0 to −12.9, p = 0.022). GAHT duration and age were not significant (p > 0.98).

### Benjamini–Hochberg correction

After applying the Benjamini–Hochberg correction for multiple comparisons, BMI remained a significant positive predictor of CMJ peak power (*p*_adj_ = 0.01, **Figure 1**). No other predictors, including MET-minutes, GAHT duration, or age, were significant after adjustment

**Figure 1.**
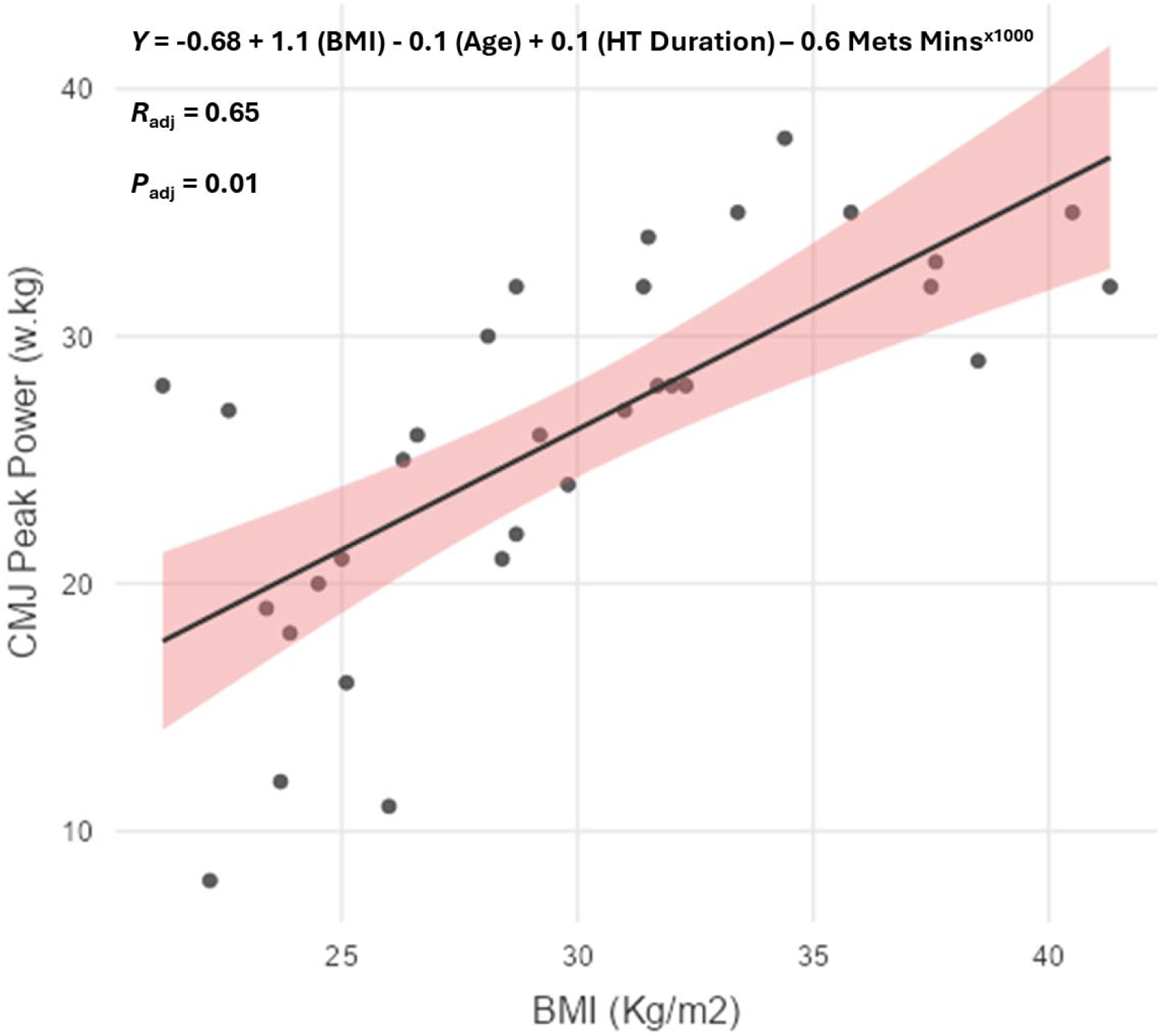
Linear Regression Analysis of Benjamini–Hochberg corrected predictors of Countermovement Jump Peak Power (PP) (W/Kg)

## Discussion

This pilot study provides the first functional characterisation of musculoskeletal health in ageing TGDW undergoing feminising HT. By combining bioimpedance-derived body composition, ultrasound-based bone health measures, and multiple indices of muscle strength and power, we offer novel insights into the musculoskeletal ageing trajectory of a population that has historically been overlooked in gerontological and clinical exercise science research.

The key findings were first, sarcopenia incidence and osteoporosis incidence were higher than in the UK population. A large majority of the participants were classified as overweight and obese (**Table 2**) also higher than the UK Population. In this cohort, only one in four TGDW participants was classified as physically active, compared with approximately two-thirds of their age-matched UK counterparts.

Finally, exploratory analyses showed that hormone therapy duration was not significantly associated with musculoskeletal outcomes after controlling for covariates, whereas BMI emerged as a stronger determinant of peak power.

The prevalence estimates suggest that ageing transgender women may face disproportionate risks of sarcopenia, mobility limitation, and osteoporosis compared with cisgender peers. These impairments are clinically meaningful, given the established links between muscle function, fall risk, disability, and mortality in older adults [34–36]. Importantly, our observation that average grip strength and lower-limb power exceeded clinical cut-offs should not obscure the high proportion of individuals already crossing diagnostic thresholds, especially as the prevalence of physically active TGDW was 64% lower than their age-matched peers.

Bone health findings reinforce concerns about skeletal vulnerability in this population. While oestrogen, a key component of feminising HT, protects by decreasing bone breakdown and stimulating bone formation, a process known as bone remodelling [37] oestrogen may not uniformly safeguard against bone loss in later life in older TGDW. Oestrogen may not consistently protect against bone loss in older TGDW, largely due to heterogeneity in therapy history, dosing regimens, and prior exposure to endogenous testosterone[38–42]. The site-specific disparities we observed, with greater vulnerability at the tibia than radius (**Table 2**), suggest uneven skeletal adaptation, although our tibia results were statistically underpowered. Our QUS precision was 2% at the Radius and 5% at the tibia (Supplementary File CV QUS [27]), and future work using more precise measures of DXA (CV 1-2%[28, 43, 44]) at the more clinically relevant sites of the total hip and lumbar spine [18] for osteoporosis and pQCT (CV 1-2% [45]) at the tibia and radius will be critical to develop and confirm these findings.

Regression analyses provided further contextualisation. BMI was a positive predictor of CMJ peak power, reflecting the established influence of greater adiposity on lower-body force generation [46]. However, excess adiposity also carries cardiometabolic risks [47] and may exacerbate mobility limitation when power is expressed relative to body mass; alternatively, relative weakness may be caused by reduced mobility, neural adaptations and changes in muscle morphology [48]. These findings challenge assumptions that feminising HT alone dictates musculoskeletal ageing trajectories. Instead, ageing transgender women appear subject to the same multifactorial influences as cisgender peers: age, adiposity, and physical activity [49], while facing a higher prevalence of adverse outcomes [7]. This dual burden highlights the urgency of extending evidence-based exercise and nutritional recommendations to ageing TGDW populations, ensuring that interventions target both sarcopenia and bone fragility alongside standard endocrinological care.

The novelty of this study lies in being, to our knowledge, the first study to attempt characterisation of musculoskeletal function and bone health in ageing TGDW. Previous research showed a focus on younger cohorts [50–54] or athletic populations [55–57], leaving older adults unexamined. By establishing prevalence estimates for sarcopenia, mobility limitation, and osteoporosis, our findings fill a critical knowledge gap and raise the possibility of health inequalities in this community.

## Limitations

Recruitment challenges for this transgender and gender diverse men compared with TGDW may reflect demographic differences, community engagement patterns, or event-specific factors. This limitation should be considered when interpreting the findings, as the findings only relate to feminising HT, and future research should aim to address this gap for masculinising HT. Several further limitations warrant caution. Recruitment from community festivals may have favoured healthier and more socially connected individuals [58], potentially underestimating the true prevalence of musculoskeletal deficits. The cross-sectional design precludes causal inference, and the modest sample size (*n* = 48) limits generalisability. Bioimpedance and ultrasound, though pragmatic in field settings, lack the precision of dual-energy X-ray absorptiometry, magnetic resonance imaging, or peripheral quantitative computed tomography. In addition, force plate measures of jump performance were collected at only one site due to inclement weather, resulting in a reduced sample size for these outcomes. Finally, comparisons with population prevalence rely on published data, which differ in methodology and case definitions.

## Future Directions

Future studies should adopt longitudinal designs in ageing TDGA undergoing with cisgender comparator groups and advanced imaging, including clinical measures such as dual-energy X-ray absorptiometry to triangulate body composition and bone health with the effects of transition and ageing. Intervention trials will be needed in the future to determine the efficacy of targeted training in reducing sarcopenia and osteoporosis risk among ageing TGDW on feminising HT. Qualitative research could also enrich our understanding by situating these physiological findings within the lived experiences of ageing, health, and identity, which we did not touch on in this study.

To strengthen statistical power in future studies measuring tibial T-scores and physical activity (**Table 3**), future research should prioritise methodological rigour over altering participant characteristics. Larger sample sizes in this cohort remain the most effective way to reduce error and detect meaningful differences. Power can be further improved through standardised protocols and high-quality measurements (e.g., peripheral quantitative computed tomography for tibia T-scores and validated accelerometery for physical activity), while adopting strict control of key confounders, such as age, type of HT, and BMI. These refinements will produce adequately powered (≥0.80), reliable, and generalisable findings.

## Conclusion

Musculoskeletal assessment in ageing TGDA populations is feasible, though functional limitations are evident in a subset of individuals. Elevated BMI, high prevalence of sarcopenia, osteoporosis, low physical activity levels, and substantial within-group heterogeneity highlight important clinical concerns. These findings emphasise the need for larger, longitudinal studies employing robust clinical measures to better characterise musculoskeletal ageing and to inform tailored screening and intervention strategies for this population.

## Data Availability

All data produced in the present work are contained in the manuscript

## Contributions

Conceptualisation, BRH; methodology, BRH and AI; writing-original draft preparation, BRH; Data collection. BRH, NM, CEL, KH, ME and AI; Data cleaning and Preparation, KH and BRH; Data analysis, BRH; writing review and editing, ALL.

## Ethics and Reporting Statement

Authors should declare at submission that all relevant ethical guidelines have been followed, all necessary IRB and/or ethics committee approvals have been obtained (ETHOS ID: 77000), all necessary patient/participant consent has been obtained, and the appropriate institutional forms have been archived.

## Acknowledgements

The authors wish to thank the BarPop™ team on Canal Street, Manchester, for generously hosting us to test during the Sparkle Festival, and the authors also extend their gratitude to Sparkle and TransPride Brighton for granting permission to conduct sampling at their respective festivals.

